# Genomic surveillance of Lassa virus in Guinea through in-country sequencing

**DOI:** 10.64898/2026.03.04.26347418

**Authors:** Jacob Camara, Giuditta Annibaldis, Joon Klaps, Kékoura Ifono, Fara Raymond Koundouno, Youssouf Sidibé, Sarah Ryter, Moussa Conde, Saa Lucien Millimono, Mette Hinrichs, Julia Hinzmann, Nils Peter Petersen, Mia Le, Annick Renevey, Ehizojie Ehiremen Emua, Philippe Lemey, Simon Dellicour, Stephan Günther, N’Faly Magassouba, Sophie Duraffour, Liana Eleni Kafetzopoulou, Sanaba Boumbaly

## Abstract

Strengthening in-country sequencing capacity generated 28 Lassa virus genomes from human clinical cases, expanding our knowledge of Lassa fever in Guinea. Phylogeographic analysis revealed cross-border exchange between Liberia and the N’Zérékoré region, and a Sierra Leone introduction into the Guéckédou area. Enhanced genomic surveillance is crucial to guide future public health actions.

## Research letter

Lassa fever is a life-threatening viral hemorrhagic disease endemic in West Africa, with early clinical features that overlap broadly with other febrile illnesses, complicating case detection and public health surveillance (1). The causative agent, Lassa virus (LASV), is an Old World arenavirus with a bi-segmented ambisense RNA genome (S and L segments) that exhibits distinct phylogenetic structure across its endemic geographic regions. Lineages I–III and VI circulate in Nigeria; lineage IV predominates across the Mano River Union countries (Guinea, Sierra Leone and Liberia); and additional distinct lineages have been identified in Mali/Côte d’Ivoire (Lineage V) and Togo (Lineage VII). Although Lassa fever cases are only reported sporadically in Guinea, serological evidence from the south-eastern (forested) and central regions indicates broad population exposure (2, 3). Sequencing efforts thus far have generated partial LASV genomes from rodent reservoirs in Upper Guinea (4), while genomes from human infections remain limited, with the 2019 Kissidougou genome being one of the few and only recently sequenced (5, 6). Sparse genomic data limits our understanding of geo-genomic variation, local outbreak dynamics, and clinical correlations. This underscores the need for sustained and increased genomic surveillance to inform diagnostics, epidemiology, and patient management.

To enhance viral surveillance and diagnostic capabilities locally, a genomic capacity strengthening project was launched in 2021 at the *Centre de recherche en Virologie - Laboratoire des Fièvres Hémorragiques Virales de Guinée* (CRV-LFHVG; Conakry, Guinea). Sequencing infrastructure was initially established in response to the SARS-CoV-2 pandemic and included targeted sequencing using Nanopore sequencing (Oxford Nanopore Technology) (7). In 2022-2023 the laboratory expanded its focus to include metagenomic sequencing approaches for virus identification (8, 9). This untargeted genomic capacity component was integrated within the diagnostic network of three surveillance laboratories for viral hemorrhagic fevers (VHFs) across Guinea: [1] the national reference laboratory in Conakry (CRV-LFHVG) and two satellite laboratories in the forest region, respectively located in [2] Guéckédou (*Laboratoire des Fièvres Hémorragiques Virales de Guéckédou* or LFHV-GKD) and [3] N’Zérékoré (*Laboratoire des Fièvres Hémorragiques Virales de Hôpital Régional de N’Zérékoré* or LFHV-HRNZE). Between 2020 and 2024 this laboratory network confirmed a total of 36 Lassa fever cases (10), including a nosocomial outbreak in Conakry in 2022. In order to investigate virus diversity in the Lassa fever confirmed cases, in-country metagenomic nanopore sequencing was performed at CRV-LFHVG. A total of 28 Lassa fever cases were successfully sequenced, all of which yielded sufficient genomic coverage for downstream phylogenetic analysis (genomic coverage varied between 36-100% per segment, Table S1). The majority yielded near-complete genomes for both segments (>90% coverage; 20/28 S segments, 18/28 L Segments) and all sequences were phylogenetically classified as lineage IV.

Phylogenetic analysis revealed that many of the newly sequenced Guinean strains are substantially divergent from previously characterized cases, with branch lengths suggesting years to decades of virus circulation in the natural reservoir prior to sampling in human cases (Figures 1, 2). Bayesian phylogeographic reconstruction estimated substitution mean rates of 8.5×10□□ (S segment) and 8.2×10□□(L segment) substitutions/site/year, placing the root of lineage IV in the 17th-18th centuries, likely originating in south-eastern (forested) Guinea (Figures 1, 2).

**Figure 1:**
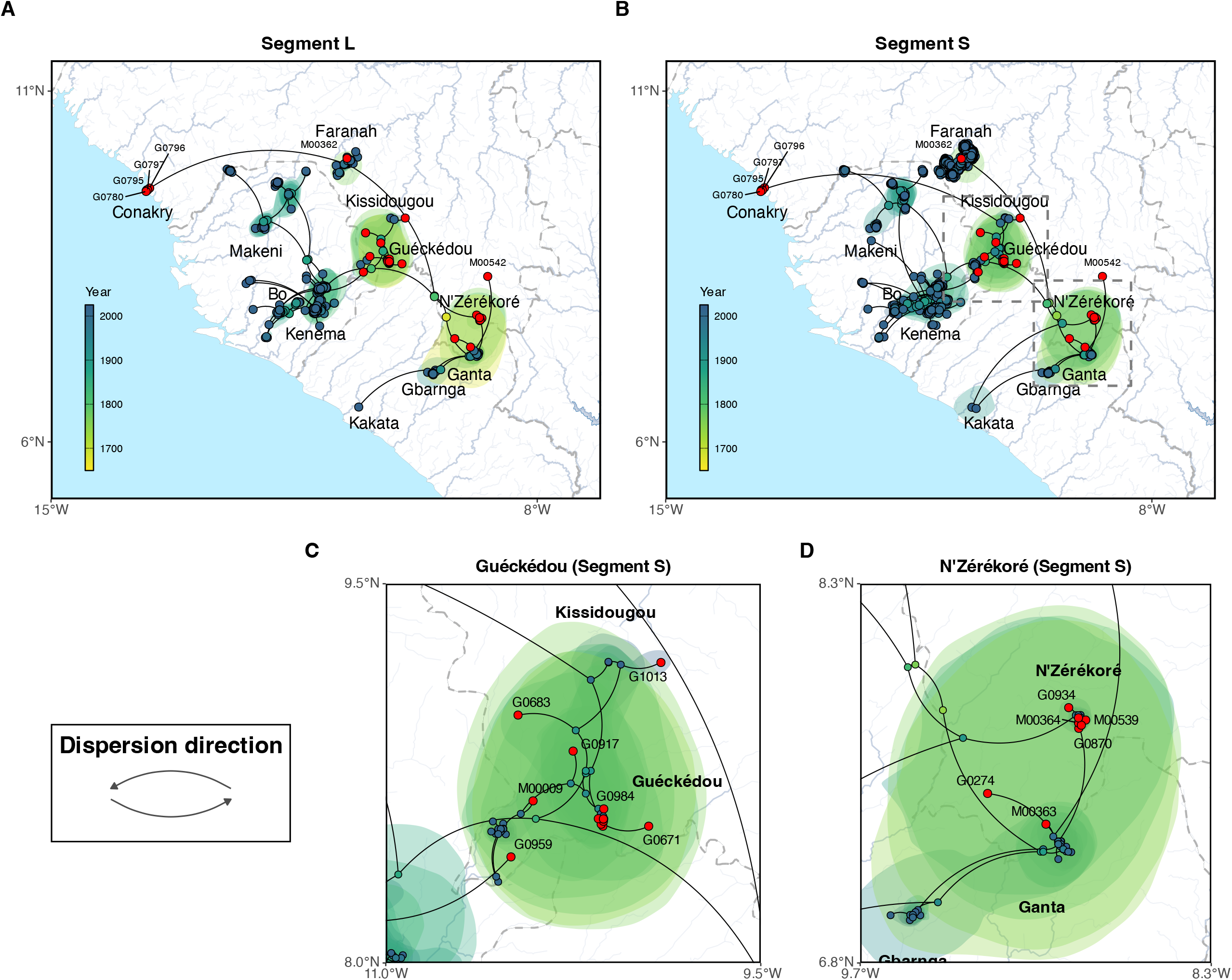
Phylogeographic reconstruction of the dispersal history of Lassa virus lineage IV. We here report the result of the continuous phylogeographic inference based on L (A) and S (B) segment sequences. For each analysis, we map the corresponding maximum clade credibility tree with internal and tip nodes colored according to their estimated time of occurrence and sampling date, respectively. Tip nodes corresponding to newly sequenced cases from this study are highlighted in red. Shaded polygons represent the 80% highest posterior density (HPD) regions, reflecting uncertainty in internal node location inference. The estimated root of lineage IV, dating to the 17th-18th centuries, is indicated by a dashed circle in the south-eastern (forested) region of Guinea. Zoomed map insets highlight specific transmission dynamics in (C) Guéckédou and (D) N’Zérékoré, illustrating the dense local clustering and the cross-border introductions from Liberia (Ganta) into the N’Zérékoré region.

**Figure 2:**
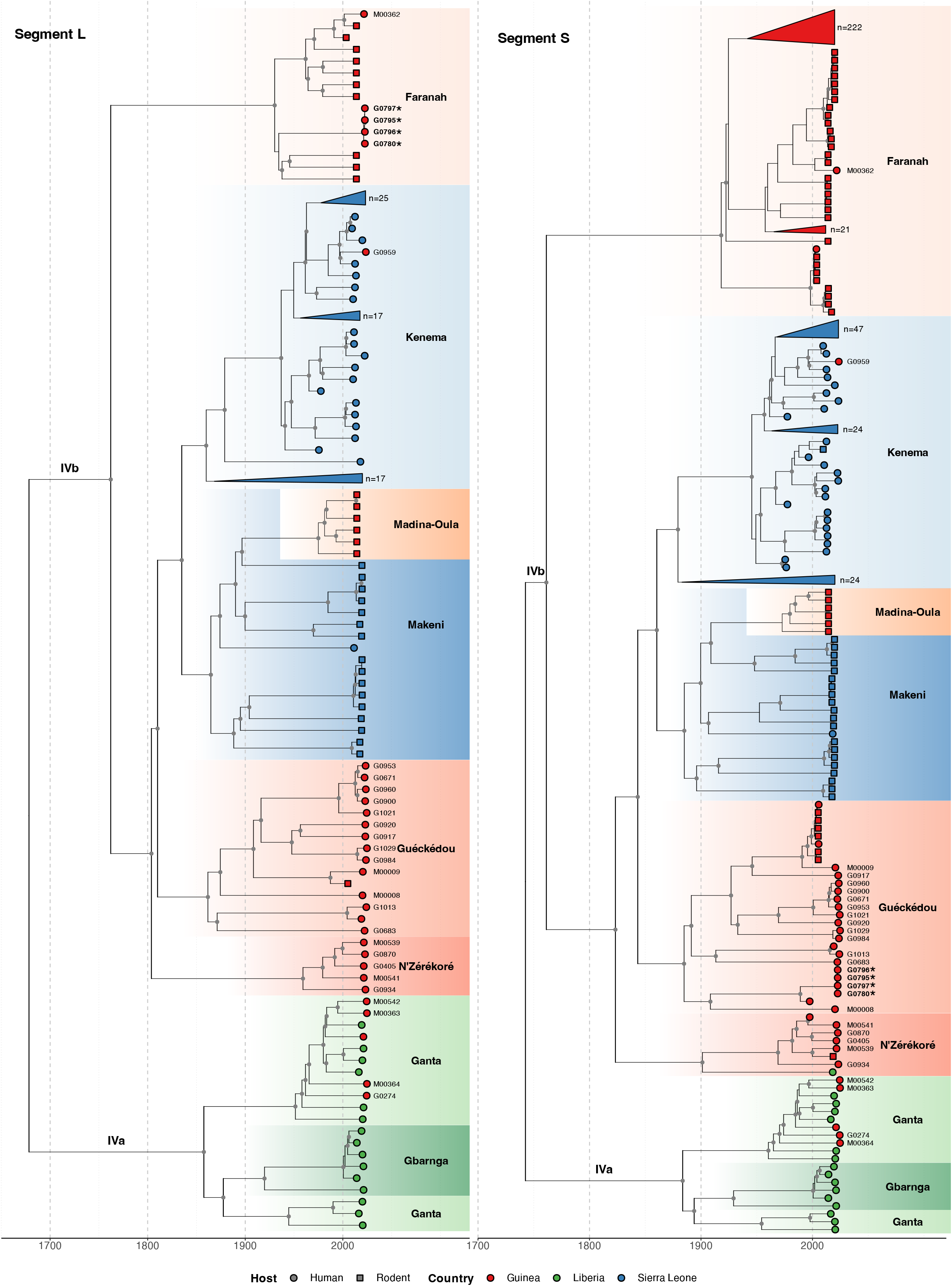
Temporal Evolution of the L and S Segments of Lassa virus in Guinea. Time scaled Maximum Clade Credibility trees for the L-segment and S-segment. Tips are colored by country of origin. Sub-lineages are annotated and colored by their predominant geographic location (e.g., Kenema, Faranah, Ganta). Clades that fall outside the sequence diversity of interest (i.e. have no association with the sequences reported in this manuscript) are collapsed and annotated with the total number of sequences they include. Internal nodes with a clade credibility of over 80% have a grey dot. Sequences reported in this manuscript have their sample identification codes indicated next to their respective tip. Two co-circulating sub-lineages of Lassa virus were detected in N’Zérékoré: a locally established IVb lineage (M00539, M00541, G0405, G0870, G0934), and the IVa lineage linked to the Liberian region of Ganta (M00363, M00542, M00364, G0274). The sample from Guéckédou (G0959) grouped within the Kenema, Sierra Leone, Lassa virus cluster, reflecting its travel-linked origin. Strains that form the nosocomial transmission chain with reassorted genomes have been highlighted in bold and have a star [*].

Spatial viral diversity in Guinea is organized into three predominant geographic clusters, mainly associated with the areas of Guéckédou, N’Zérékoré, and Faranah. This structure should be interpreted cautiously, as Guéckédou and N’Zérékoré both have surveillance laboratories, and sequences from the Faranah region originate from rodent reservoirs sampled during previous studies. Consequently, the inferred geographic clustering may be driven, at least in part, by uneven sampling intensity. Two co-circulating sub-lineages of Lassa virus were identified in the N’Zérékoré region: [1] an older IVb lineage, which has been long-established locally (M00539, M00541, G0405, G0870, G0934), and [2] the IVa lineage, which traces back to multiple independent introductions (M00363, M00542, M00364, G0274) from the Ganta area located in north-eastern Liberia with common ancestors dating to the 1950s - 1980s (Figures 1D, 2). In Guéckédou, we detected an imported Lassa virus infected case originating from the Kenema area of Sierra Leone (G0959, Figures 1C, 2), illustrating that sporadic introductions from neighboring endemic regions to Guinea can occur and contribute to Guinean Lassa virus diversity.

Four Lassa fever cases were sequenced from Conakry (capital city, western part of Guinea), all of which were associated to a previously identified nosocomial transmission chain linked to a travel case from an endemic area. The Lassa virus sequences recovered (G0780, G0795, G0796, G0797) show minimal between sequence variation (0 - 4 mutations in L segment, 0 - 1 mutations in S segment), consistent with a single transmission event. Furthermore, all four genomes were identified as a reassortant LASV variant with their L and S segments clustering significantly differently (supplementary methods). Their S segments cluster with sequences previously identified in the Guéckédou region (Figure 2, Lineage IVb, sequences marked with a star [*]; Guéckédou sequence grouping) and their L segments with sequences previously identified in the Faranah region (Figure 2, Lineage IVb, sequences marked with a star [*]; Faranah cluster).

This study increases the available Lassa virus sequences derived from human clinical cases and provides new genomic insights into Lassa virus circulation in Guinea. These findings were made possible through strengthened laboratory diagnostics in Lassa fever endemic areas (Guéckédou and N’Zérékoré), and the establishment of new sequencing capacity at the national reference laboratory for viral hemorrhagic fevers in Conakry. Ongoing monitoring and genomic surveillance remain crucial for guiding public health interventions as well as the development of medical counter measurements, including vaccines, monoclonal antibody therapies and rapid diagnostic tests.

## Data Availability

All data produced in the present study are available upon reasonable request to the authors

## Contributions

Conceived and designed the study: G.A., F.R.K., Y.S., S.G., N.F.M., S.Du., L.E.K., S.B.

Collected data and/or performed laboratory diagnostics: J.C., G.A., K.I., F.R.K., Y.S., S.R., M.C., S.L.M., M.H., J.H.

Performed sequencing and/or sequence validation: J.C., G.A., K.I., S.R., M.C., S.L.M., M.H., J.H., N.P.P., M.L., A.R.

Formal phylogenetic analysis: J.C., G.A., J.K., K.I., S.R., M.C., S.L.M., S.De.

Phylogeography analysis: J.K., S.De.

Project implementation: J.C., G.A., J.K., K.I., F.R.K., Y.S., S.R., M.H., J.H., A.R., E.E.E., P.L., S.G., N.F.M., S.Du., L.E.K., S.B.

Funding acquisition: P.L., S.G., N.F.M., S.Du., S.B.

Wrote the manuscript: J.C., G.A., J.K., K.I., S.G., N.F.M., S.Du., L.E.K., S.B.

Edited the manuscript: all authors.

All authors read and approved the contents of the manuscript.

## Acknowledgments

The authors thank the Agence Nationale de Sécurité Sanitaire (ANSS), the Ministry of Health of the Republic of Guinea, the “Délégations Régionale et Préfectorale de la Santé” and the healthcare workers involved in the response.

## Declaration of generative AI and AI-assisted technologies in the writing process

During the preparation of this work the author(s) used ChatGPT / free version in order to edit some sentences. After using this tool/service, the author(s) reviewed and edited the content as needed and take(s) full responsibility for the content of the publication.

## About the Authors

Mr. Camara is leading the sequencing team at Centre de recherche en Virologie - Laboratoire des Fièvres Hémorragiques Virales de Guinée (CRV-LFHVG) since 2023, where he oversees genomic sequencing activities and data generation.

Dr. Annibaldis is based at the Bernhard Nocht Institute for Tropical Medicine (BNITM) in Hamburg, where she coordinates strengthening projects for viral hemorrhagic fevers (VHFs) surveillance. Her work focuses on sequencing capacity and laboratory system strengthening in Guinea and Nigeria.

Mr. Klaps is working as a PhD student in the laboratory of Evolutionary and Computational Virology at the Rega Institute (KU Leuven, Belgium). His work focuses on expanding the computational infrastructure for virological surveillance and intra-host sequencing analyses.

Mr. Ifono has worked at the Laboratoire des Fièvres Hémorragiques Virales de Guéckédou (LFHV-GKD) since 2017 and currently serves as Deputy Lead for diagnostic VHF surveillance in the same laboratory. From 2021 to 2022, he led the sequencing unit at CRV-LFHVG in Conakry.

## Supplementary files

### Materials and methods

#### Ethical approval and Nagoya permit

This descriptive research, using anonymized diagnostic surveillance data, has been approved by the National Ethics Committee of Guinea (CNERS) under the number 009/CNERS/25. This work is part of the Nagoya permit number 006/2023/PN.

#### Laboratory investigations

EDTA-blood was collected from VHFs suspected cases. Real-time reverse transcription polymerase chain reaction (RT-PCR) was performed at the *Centre de recherche en Virologie - Laboratoire des Fièvres Hémorragiques Virales de Guinée* (CRV-LFHVG), the *Laboratoire des Fièvres Hémorragiques Virales de Guéckédou* (LFHV-GKD) and *Laboratoire des Fièvres Hémorragiques Virales de Hôpital Régional de N’Zérékoré* (LFHV-HRNZE). The detailed description of laboratory surveillance for Lassa fever in Guinea and Lassa cases is provided elsewhere (*Annibaldis et al*., *manuscript submitted for publication*) (1). Viral RNA was extracted with the QIAamp viral RNA extraction kit (Qiagen, Germany) using 70 μl of the human plasma (or serum) together with 70 µl of nuclease-free water and processed according to the manufacturer’s instructions with the addition of two rounds of buffer AW2 washes and a 10-minute dry spin. RNA extracts were used for RT-PCR on the Rotor-Gene Q platform (Qiagen). Leftover RNAs were stored at −20°C. For Lassa virus diagnostics, the RealStar® Lassa Virus RT-PCR Kit 2.0 from altona Diagnostics (Germany) was used. A Lassa virus positive RT-PCR result is defined as at least one of the two targets (S or L-segment) being detected as per manufacturer instructions.

#### Metagenomic sequencing

Leftover RNA extracts or newly extracted RNAs were used for nanopore next generation sequencing on the MinION platform (Oxford Nanopore Technologies (ONT), United Kingdom). RNA extracts and sequencing libraries were prepared as described previously at CRV-LFHVG (2-4). Briefly, viral RNA was digested with DNase (TURBO DNase, Thermo Fisher Scientific) and then randomly reverse-transcribed, and amplified using a Sequence Independent Single Primer Amplification (SISPA) approach. MinION sequencing libraries were prepared using the Ligation Sequencing Kit (SQK-LSK109) according to manufacturer’s instructions. Libraries were loaded onto the R9.4.1 Flow Cells (FLO-MIN106D, ONT) and run on the Mk1C (ONT) device. Sequencing flow cells were reloaded with leftover libraries after 24 hr. Runs were further stopped after ∼48 hr and fast5 files were transferred to a laptop for basecalling and demultiplexing. Before 2025, a fastq files were generated from the fast5 files using Guppy v5.0.16, and consensus genomes were obtained using minimap2 v.2.17 and CANU v1.9 (3). For this work, consensus genomes were re-generated using Dorado v0.7.2 and the upgraded metagenomic nanopore pipeline ViMOP (5). The majority consensus sequence consisted of bases called at a minimum depth of 20x and 70% base predominance per nucleotide location. The complete sequences have been submitted to GenBank (GenBank IDs: PV847661-68; PX115263-PX115312).

#### Phylogenetic analysis

All publicly available Lassa virus (LASV) sequences were downloaded from NCBI Virus GenBank on July 25, 2025 (*Mammarenavirus lassaense* species, taxid:3052310). Sequence deduplication was performed using MMseqs2 v14.7e284 (6) to identify duplicates based on high sequence similarity (≥99.9% identity based on high sequence similarity (≥99.9% identity and <2 mismatches). Sequences shorter than 500 bp, sequences associated with patents or vaccines, and sequences lacking sufficient geographic precision were excluded. Only sequences from Guinea, Sierra Leone and Liberia were kept. Genes were extracted and aligned to reference sequences using MAFFT v7.508 (7), then concatenated in a consistent orientation allowing codon partitioning. To identify potential outliers and validate the alignment, a maximum likelihood tree was constructed using IQ-TREE v2.1.4 (8). Misaligned regions were manually inspected and corrected.

Time-calibrated phylogenies were estimated in BEAST X v10.5.0 (9), with a chain of 2 x 500 billion interactions using a GTR+*Γ* 4 substitution model with codon partitions, an uncorrelated lognormal relaxed molecular clock, and a Bayesian skygrid coalescent prior for population dynamics. Spatial diffusion was modelled with a Relaxed Random Walk model. Logs were combined after discarding the first 10 million iterations of each run as burn-in, and posterior trees were subsequently summarized as maximum clade credibility (MCC) trees using TreeAnnotater v10.5.0 (9). Visualization of the spatiotemporal dispersal history was performed using the ‘ggphylogeo’ v.0.1.2 and ‘ggtree’ v4.0.4 R packages (9, 10). Reassortment analysis of the concatenated L and S segments in RDP4(11), using a full exploratory recombination scan, identified samples G0796, G0797, and G0795 as significant recombinants. These events were strongly supported by five out of nine algorithms: Bootscan (p = 1.65E-03), Maxchi (p = 4.70E-12), Chimaera (p =3.33E-07), SiSscan (p = 4.42E-50) and 3Seq (p = 2.83E-19), but were not significant by RDP, GENECONV, PhyloPro and LARD (12-20).

---

**Supplementary Table 1.**

| Nr | Seq ID | Location | Collection period | Ct value S diag | Ct value L diag | Coverage S [%] | Coverage L [%] | GenBank ID S | GenBank ID L |
| --- | --- | --- | --- | --- | --- | --- | --- | --- | --- |
| 1 | M00008 | Guéckédou | Mar-2020 | 36.2 | 31.3 | 72.9 | 60.8 | PX115298 | PX115297 |
| 2 | M00009 | Guéckédou | Jul-2020 | 25.6 | 21.7 | 99.8 | 99.6 | PX115300 | PX115299 |
| 3 | G0274 | Yomou | May-2021 | 29.6 | 29.5 | 91.6 | 84.6 | PX115264 | PX115263 |
| 4 | M00542* | Beyla | May-2021 | 40.4 | 33.4 | 77.9 | 62.9 | PX115311 | PX115312 |
| 5 | G0405 | N'Zérékoré | Jun-2021 | 28.7 | 26.5 | 99.3 | 99.4 | PX115266 | PX115265 |
| 6 | M00541 | N'Zérékoré | Jun-2021 | 31.2 | 34.8 | 88.6 | 88.9 | PX115310 | PX115309 |
| 7 | M00539 | N'Zérékoré | Jul-2021 | 27.3 | 45.0 | 99..8 | 99..3 | PX115307 | PX115308 |
| 8 | M00363 | Yomou | Aug-2021 | 23.4 | 22.0 | 99.8 | 99.6 | PX115304 | PX115303 |
| 9 | M00364 | N'Zérékoré | Aug-2021 | 32.7 | 26.5 | 96.0 | 98.1 | PX115306 | PX115305 |
| 10 | M00362 | Faranah | Sep-2021 | 21.3 | 19.3 | 99.4 | 99.7 | PX115302 | PX115301 |
| 11 | G0683 | Guéckédou | Apr-2022 | 33.9 | 30.5 | 62.2 | 69.8 | PX115270 | PX115269 |
| 12 | G0671 | Guéckédou | Apr-2022 | 32.3 | 29.0 | 70.6 | 70.9 | PX115267 | PX115268 |
| 13 | G0780** | Conakry | Aug-2022 | 30.0 | 31.2 | 35.7 | 42.3 | PX115272 | PX115271 |
| 14 | G0795** | Conakry | Aug-2022 | 26.0 | 23.5 | 95.5 | 94.7 | PV847661 | PV847662 |
| 15 | G0796** | Conakry | Aug-2022 | 23.3 | 21.0 | 98.3 | 99.7 | PV847663 | PV847664 |
| 16 | G0797** | Conakry | Aug-2022 | 21.4 | 23.7 | 95.6 | 99.4 | PV847665 | PV847666 |
| 17 | G0870 | N'Zérékoré | Sep-2022 | 31.6 | 28.1 | 43.3 | 67.0 | PX115273 | PX115274 |
| 18 | G0900 | Guéckédou | Dec-2022 | 21.7 | 23.5 | 99.8 | 99.6 | PX115276 | PX115275 |
| 19 | G0917 | Guéckédou | Jan-2023 | 17.7 | 18.0 | 99.9 | 99.6 | PX115278 | PX115277 |
| 20 | G0920 | Guéckédou | Jan-2023 | 30.6 | 27.4 | 72.2 | 62.3 | PX115279 | PX115280 |
| 21 | G0934 | N'Zérékoré | Mar-2023 | 28.0 | 24.2 | 98.2 | 43.7 | PX115282 | PX115281 |
| 22 | G0953 | Guéckédou | May-2023 | 23.7 | 19.3 | 99.6 | 99.6 | PX115283 | PX115284 |
| 23 | G0959 | Guéckédou | Jul-2023 | 30.6 | 20.2 | 99.6 | 95.4 | PX115286 | PX115285 |
| 24 | G0960 | Guéckédou | Aug-2023 | 45.0 | 25.9 | 96.0 | 93.3 | PX115287 | PX115288 |
| 25 | G0984 | Guéckédou | Sep-2023 | 22.6 | 24.7 | 99.6 | 99.5 | PX115289 | PX115290 |
| 26 | G1013 | Kissidougou | Jan-2024 | 18.4 | 19.3 | 94.5 | 97.7 | PX115292 | PX115291 |
| 27 | G1021 | Guéckédou | Jun-2024 | 20.5 | 19.5 | 98.9 | 98.6 | PX115294 | PX115293 |
| 28 | G1029 | Guéckédou | Sep-2024 | 14.5 | 16.2 | 99.5 | 97.3 | PX115296 | PX115295 |

\*Confirmed LF case detected after retrospective testing
\*\*Cases from the nosocomial LF outbreak in Conakry, 2022

